# Federated learning in a regulator-audited secure processing environment: a multi-hospital deployment study

**DOI:** 10.64898/2026.09.15.26363093

**Authors:** Valtteri Nieminen, Hartmut Schultze, Sampo Kukkurainen, Harri J. Rantala, Leena Hakkarainen, Per-Eric Gustafsson, Tuomas Hakala, Emmi Turunen, Markus Johansson, Tarja Laitinen, Dimitrios Tsallos, Kimmo Porkka, Arho Virkki, Joachim Schultze, Eric Fey

**Affiliations:** Helsinki University Hospital, Helsinki, Finland; Department of Computing, University of Turku, Turku, Finland; German Center for Neurodegenerative Diseases (DZNE), Bonn, Germany; 42Hills GmbH, Königswinter, Germany; Tampere University Hospital, Tampere, Finland; Auria Clinical Informatics, Turku University Hospital, Turku, Finland; Tieto Oyj, Espoo, Finland; LUT University, Lappeenranta, Finland; Istekki Oy, Kuopio, Finland; Institute for Molecular Medicine Finland FIMM, HiLife, University of Helsinki, Helsinki, Finland; iCAN Digital Precision Cancer Medicine Flagship, University of Helsinki and Helsinki University Hospital, Helsinki, Finland; Department of Mathematics and Statistics, University of Turku, Turku, Finland; Genomics and Immunoregulation, Life and Medical Sciences (LIMES) Institute, University of Bonn, Bonn, Germany

**Author notes:** contributed equally (Valtteri Nieminen, Hartmut Schultze).

## Abstract

**Background:** Federated learning enables collaborative model development without transferring patient-level data between institutions. However, translation into routine healthcare practice remains limited because deployment requires more than distributed model training. Federated infrastructures must operate within secure processing environments, satisfy governance and security requirements, support auditability, and integrate with existing regulatory workflows for secondary use of health data. Although federated learning has been demonstrated in multiple clinical applications, evidence for its deployment, assessment, and operation within regulated health-data infrastructures remains limited.

**Methods:** We implemented a decentralised swarm learning system for federated learning and integrated it into Acamedic, a certified secure processing environment (SPE) operated by Helsinki University Hospital (HUS). Introduction of the swarm learning coordination layer constituted a material modification to the previously certified HUS SPE and triggered a differential regulatory security assessment under Finland’s Act on the Secondary Use of Health and Social Data and associated Findata requirements. The assessment evaluated controls relating to data isolation and locality, identity and access management, logging and monitoring, network security, and environment protection, while establishing a governance model that separates certification of infrastructure-level controls from study-specific assessment of data, models, parameter exchanges, and outputs. The swarm learning architecture was developed and deployed across the university hospitals of Helsinki, Turku, and Tampere, with HUS serving as the regulator-assessed implementation within a certified SPE and partner sites operating under local institutional governance frameworks. As an operational exemplar, we trained federated DeepSurv survival models for acute myeloid leukaemia (AML) using harmonised longitudinal laboratory data.

**Findings:** The differential security assessment identified one high-severity, two medium-severity, and two low-severity findings. All high- and medium-severity findings were remediated and verified, after which an independent certification report was issued for the assessed extension. The resulting federated-learning capability was authorised for secondary use of health data and operationalised as a reusable extension to the HUS secure processing environment. Swarm learning was successfully executed across three university hospitals without transferring patient-level data, achieving 100% parameter-merge success. Training operations were traceable through infrastructure logs, container logs, and a distributed-ledger coordination layer recording participant registration, parameter exchanges, and model-training lineage. On independent test sets from all three hospitals, the swarm-trained AML model demonstrated stronger risk stratification than locally trained reference models, with consistently stronger risk separation, higher log-hazard ratios (swarm vs local: 4·32 vs 1·79 at HUS, 7·20 vs 3·19 at TAYS, and 5·90 vs 2·91 at TYKS), and improved discrimination metrics. The resulting federated-learning capability was authorised for secondary use of health data, operationalised as a reusable extension to the HUS secure processing environment, and made available for future permit-approved analyses.

**Interpretation:** This study demonstrates that federated learning can be integrated into a certified secure processing environment, subjected to regulatory assessment, and operated across independent hospitals without centralising patient-level data. The principal contribution is a reusable governance and infrastructure model that separates assessment of system-level controls from study-specific evaluation of data, models, parameter exchanges, and outputs. By enabling federated learning to function as an assessed infrastructure capability rather than a project-specific exception, the approach provides a practical pathway for operationalising decentralised federated learning within regulated health-data environments. The findings are directly relevant to emerging European frameworks for secondary use of health data, including secure processing environments envisioned under the European Health Data Space, whose implementation builds on many of the same governance principles evaluated in this study.

## Introduction

Artificial intelligence (AI) has substantial potential to improve diagnosis, prognostication, treatment selection, and health-system efficiency.^1^ Yet clinical AI remains highly dependent on access to large, representative datasets for model development and validation.^2^ In practice, such datasets are rarely available within a single hospital, and conventional multi-centre AI studies have relied on centralising patient-level data under data-sharing agreements.^3^ This model is increasingly difficult to scale because health data are legally protected, operationally sensitive, and subject to strict institutional and legal governance.^4–6^

The concept of analysing health data across institutions without centralising patient-level data predates federated learning. Large distributed research networks such as OHDSI and the European DARWIN EU initiative have demonstrated the value of federated analytics, whereby institutions retain local control of data while contributing aggregate results to distributed analyses.^7,8^ However, these approaches are primarily designed for population-level analyses and distributed statistical analyses rather than collaborative development of modern AI models. Federated learning (FL) extends this paradigm by enabling institutions to collaboratively train models while data remain local, exchanging model updates rather than analytical results.^9^ Swarm learning extends this concept through a decentralised, peer-to-peer architecture that combines edge computing with blockchain-based coordination and removes the need for a central coordinator.^10^ However, translating federated learning from a research methodology into an operational healthcare capability requires more than model training.

Despite this promise, most medical FL studies remain technical demonstrations, simulations of federated settings, or analyses based on public or retrospectively partitioned datasets.^11,12^ Real-world deployments across independent healthcare institutions remain uncommon, and only a limited number of studies have reported FL or related decentralised learning workflows implemented in operational clinical environments.^9,13–15^ This implementation gap is important because deployment in hospitals requires more than a working algorithm: it requires governance, audited infrastructure, controlled access, logging, network security, and clear accountability between participating organisations.^12^ Consequently, the primary barrier to real-world adoption is increasingly infrastructural and organisational rather than algorithmic.

In Europe, secondary use of health data is increasingly being formalised through secure processing environments (SPEs) and trusted research environments in which data access, processing, audit trails, and export controls are governed within controlled technical and organisational boundaries. These frameworks are increasingly viewed as the operational foundation for secondary use of health data under the European Health Data Space (EHDS).^5,16^ FL is conceptually aligned with this direction because data remain local, but its integration into audited clinical data infrastructures has rarely been demonstrated. Emerging frameworks such as TEHDAS2 define harmonised technical and governance requirements for cross-border health-data access and highlight the need for trustworthy coordination mechanisms between institutions.^17,18^ Swarm learning’s decentralised architecture provides one potential solution by enabling verifiable coordination across institutions without reliance on a central intermediary.^10^ In contrast to conventional federated architectures that rely on a central coordinator, swarm learning provides decentralised coordination and a shared transaction log, features that align well with multi-organisational governance requirements. Whether such architectures can be deployed within audited healthcare infrastructures while supporting clinically meaningful analyses remains an open question.

Here, we report a regulator-audited swarm learning deployment integrated into a secure processing environment across three Finnish university hospitals. The system was implemented as a federated network operating under established governance and security controls, including integration into a Findata-audited secure processing environment. To evaluate the infrastructure under real-world conditions, we conducted a federated acute myeloid leukaemia (AML) survival-modelling study as an operational exemplar. By combining decentralised learning with regulator-audited infrastructure, the system enables collaborative model development while preserving local data control, supporting compliance with national and European data-protection requirements, and providing transparent audit trails of training operations. This study provides practical evidence for the feasibility of audited multi-hospital federated learning in controlled clinical data environments and illustrates the potential of regulator-audited decentralised learning to improve clinical risk stratification beyond local-only training.

## Methods

### System architecture: integration of swarm learning into the secure processing environment

Swarm learning was implemented as an additive extension to an existing secure processing environment (SPE), preserving the baseline security and access model while enabling decentralised cross-institution model training (Figure 1). In this configuration, analytics are executed entirely within the SPE boundary, with authorised researchers executing approved analytical code under existing institutional governance procedures.

**Figure 1.**
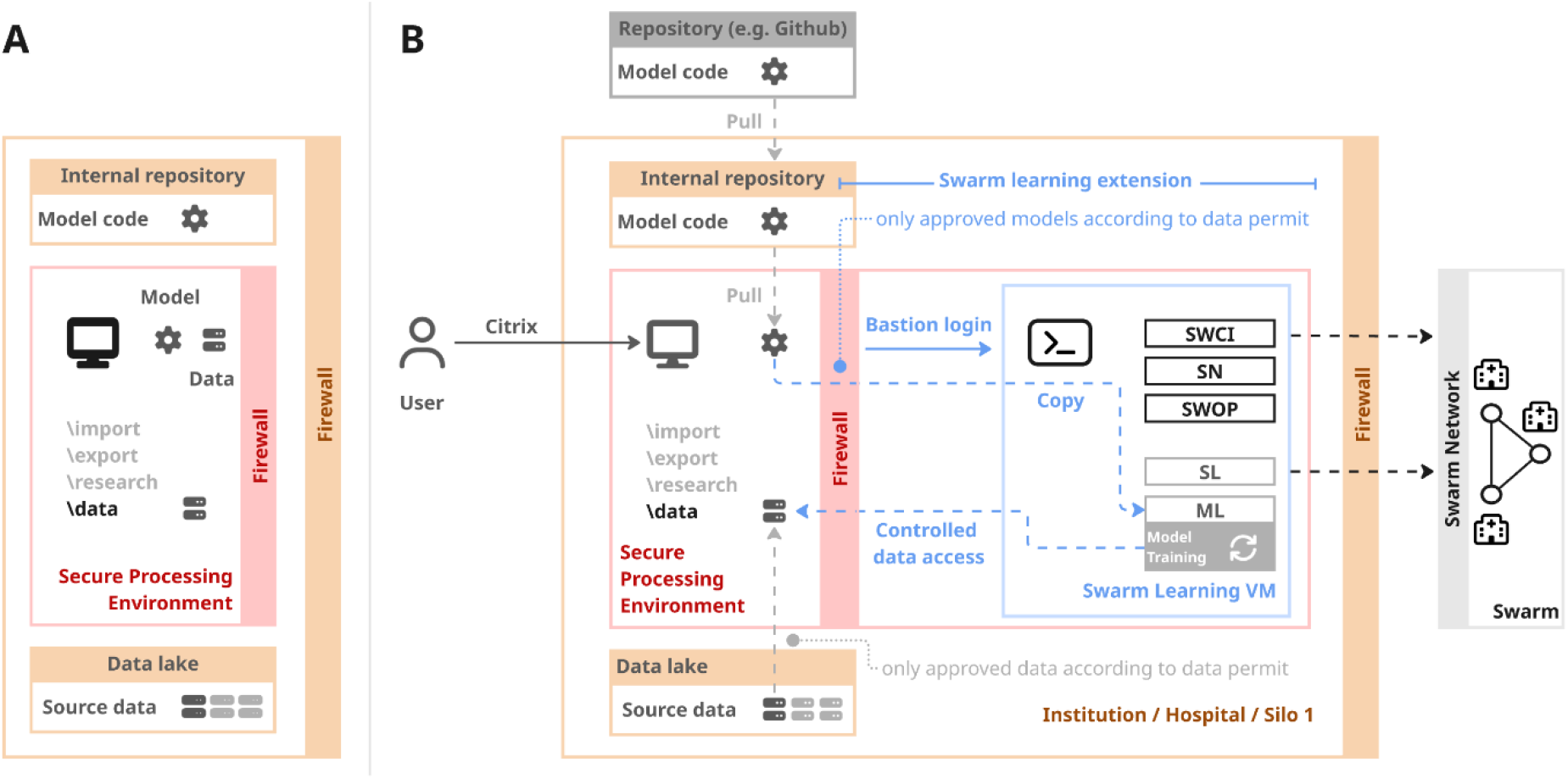
Technical architecture for integrating swarm learning into a secure processing environment. Overview of how swarm learning is introduced as an additive extension to an existing Secure Processing Environment (SPE) while preserving baseline security controls and data-access safeguards. **(A)** Baseline SPE configuration, in which authorised users access curated institutional data within the controlled environment and execute approved analytical model code; all data ingress, egress, and execution are governed by the SPE firewall and security policies. **(B)** Extended architecture enabling swarm learning within the same institutional boundary. Model code is retrieved from approved internal or external repositories and made available to a dedicated, isolated swarm learning virtual machine (VM) through controlled access mechanisms. Swarm learning components operate entirely within the SPE perimeter, training models on local data and communicating only encrypted model parameters and coordination signals with external swarm peers via restricted network interfaces. Raw data never leave the SPE, and all interactions between the SPE core, the swarm learning VM, and the external swarm network are explicitly governed by firewall rules and zero-trust access controls.

**Figure 2.**
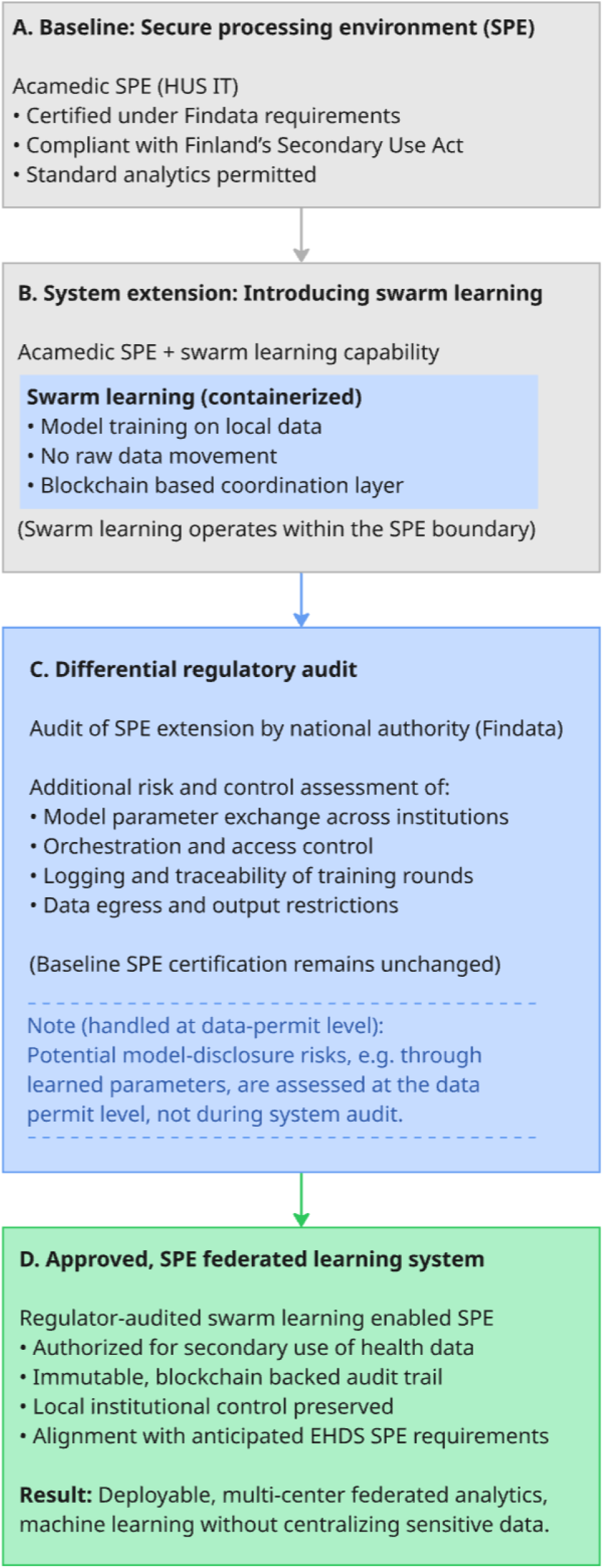
Governance and regulatory pathway for operationalising swarm learning into a secure processing environment. Overview of the staged extension of a regulator-audited Secure Processing Environment (SPE) to enable federated swarm learning under national health data legislation. **(A)** Baseline configuration of the Acamedic SPE operated by HUS IT, certified under Findata requirements and compliant with Finland’s Act on the Secondary Use of Health and Social Data, permitting standard secondary-use analytics within a controlled environment. **(B)** System extension introducing containerised swarm learning capabilities, enabling decentralised model training on local data without raw data movement, coordinated through a blockchain-based layer, while operating fully within the SPE boundary. **(C)** Differential regulatory audit conducted by the national data authority (Findata), assessing the additional risks and controls introduced by swarm learning, including cross-institution parameter exchange, orchestration and access control, logging and traceability, and data egress and output restrictions; baseline SPE certification remained unchanged. Potential disclosure risks related to trained model parameters were explicitly excluded from the system audit and are addressed at the level of individual data-permit assessments.* **(D)** Resulting regulator-audited, EHDS-ready federated learning system, authorised for secondary use of health data, preserving local institutional control and enabling multi-centre analytics without centralising sensitive patient-level data. *Assessment of risks related to potential disclosure of sensitive information through model parameters depend on specific model-data combinations and is performed at the level of individual data permits, where the proposed data–model combination and analytic outputs are reviewed, rather than during system-level auditing of the secure processing environment.

**Figure 3.**
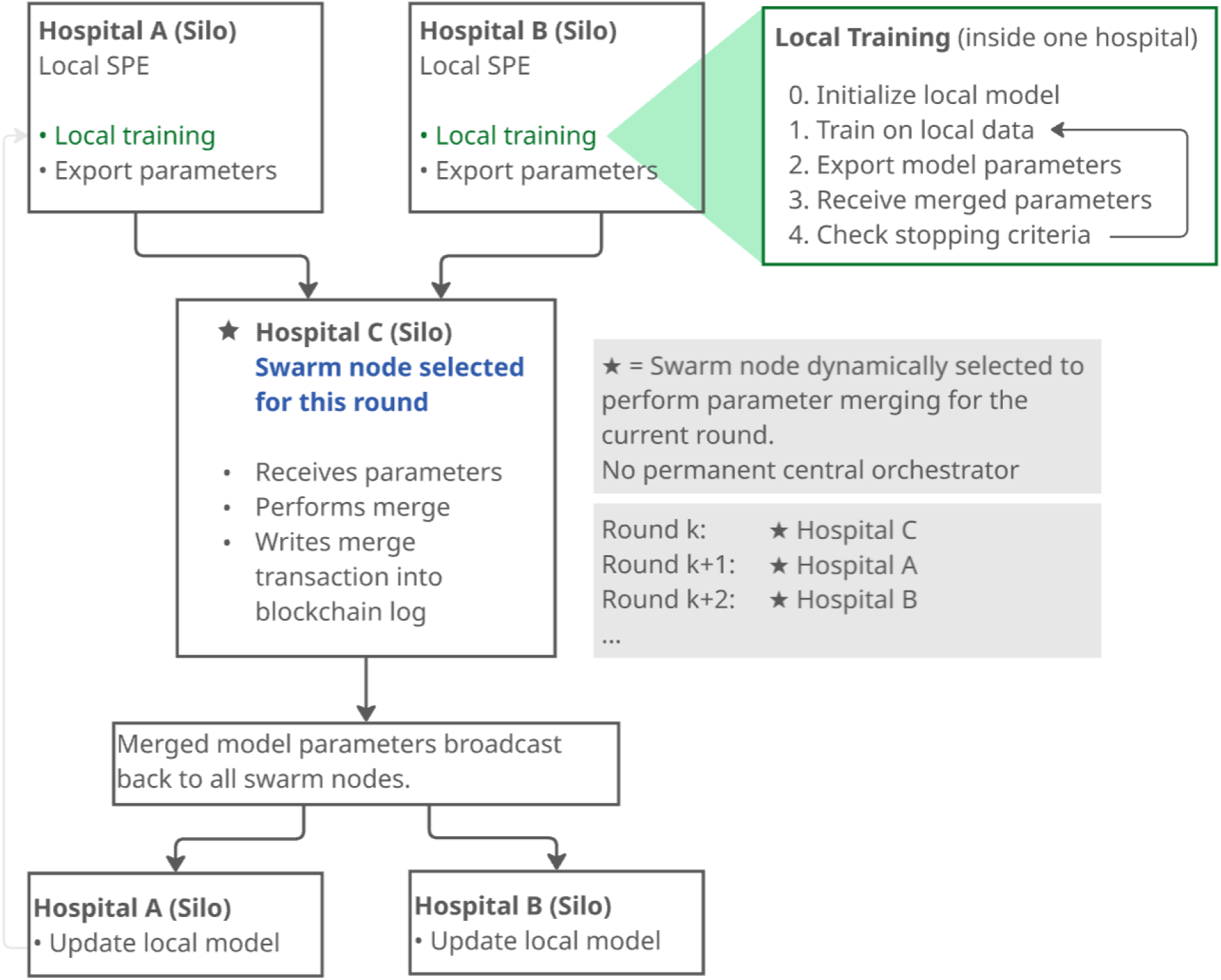
Swarm learning execution and federated training workflow. Each hospital trains a local model on site and shares only encrypted model parameters. For each training round, one swarm node is dynamically selected to perform parameter merging and to record training events on the blockchain; this role can shift between nodes across rounds. The merged model parameters are then redistributed to all participating nodes. No fixed central orchestrator exists, and raw data never leave institutional secure processing environments.

To support swarm learning, a dedicated isolated virtual machine (VM) was introduced within the institutional environment (Figure 1B). Swarm learning components operated within this logically separated execution environment and communicated through explicitly defined network pathways. Model execution remained under local institutional control, and only code approved under the applicable data permit was permitted to run within the environment.

Network connectivity for swarm learning was explicitly constrained. Participation in the external swarm network required configuration of SPE firewall rules permitting outbound communication to predefined peer endpoints, while inbound connectivity is not required, but can be configured to support more efficient peer-to-peer coordination. Where enabled, inbound access was restricted to a predefined allow-list of peer IP addresses. Within the SPE, the swarm learning virtual machine was further isolated through internal firewall controls, permitting only explicitly defined interactions with the SPE core services and preventing unrestricted lateral access.

Operational responsibilities were deliberately separated according to the principle of least privilege: swarm-learning administrators were responsible for provisioning infrastructure including provisioning the swarm learning VM and deploying the required swarm learning containers, whereas authorised researchers retained exclusive access to local data and were responsible for initiating swarm training workflows and model execution. Together, these measures established clear trust boundaries between infrastructure administration, federated coordination, and sensitive data access.

Model code and container images were introduced through established SPE governance procedures. Analytical code could be imported through approved data-ingress mechanisms; retrieved from authorised repositories or transferred through existing controlled SPE import workflows. Containerised swarm-learning components were provisioned by authorised administrators using the same governance processes applied to other software deployed within the environment. Introduction of swarm-learning functionality did not alter established procedures for software import, code review, data ingress, or result egress. All code execution remained subject to applicable data permits and institutional governance controls.

### Regulatory assessment methodology

The swarm learning functionality was evaluated as a regulated extension to an existing Findata-certified secure processing environment. The baseline secure processing environment (Acamedic, operated by HUS IT) is certified under Findata requirements and compliant with Finland’s *Act on the Secondary Use of Health and Social Data*, permitting secondary-use analytics on sensitive health data within a controlled computing boundary. Introduction of the swarm learning coordination layer constituted a material modification to the previously certified HUS Acamedic SPE and therefore triggered a differential regulatory security assessment. The assessment was limited to the HUS deployment; partner institutions implemented the same swarm learning architecture within their local processing environments and governed its use according to local institutional policies and approval procedures.

The differential assessment focused on risks introduced by federated coordination while leaving the baseline SPE certification unchanged. Assessment domains included controls for data isolation and locality and data exfiltration risk (cross-institution parameter exchange and restrictions on data egress), identity and access management (identification and user access rights governing federated participation), logging and monitoring (audit logging, traceability, and operational oversight), and environment and data protection (network isolation, encryption, and system-level data protection safeguards) (Suppl. Table 1). Particular emphasis was placed on verifying that swarm learning operations remained within the approved security perimeter and that no patient-level data were exchanged between institutions.

Swarm learning additionally incorporated a blockchain-based coordination layer that records participant registration, parameter-synchronisation events, and model-update exchanges in a distributed ledger replicated across participating sites. These records provide a persistent technical history of federated training activities and support reconstruction of model provenance, training lineage, and inter-site coordination events. Such capabilities were considered relevant to audit logging, traceability, and accountability requirements for secure processing environments.

Potential disclosure risks arising from specific data-model combinations, including risks related to trained model parameters, were not assessed at the infrastructure level.^19^ These risks were addressed through established data-permit processes governing study-specific data use, analyses, and result disclosures.

### Execution protocol for distributed training

Swarm learning experiments were performed after obtaining all required site-specific approvals and infrastructure configuration. At Helsinki University Hospital (HUS), training was conducted within the regulator-audited Acamedic SPE. Participating hospitals executed the same swarm learning workflow within their respective institutional processing environments.

Prior to execution, swarm learning infrastructure and containerised services were provisioned by designated swarm learning administrators. Authorised researchers initiated model training using approved local datasets. All data preprocessing and model execution were performed locally, and patient-level data remained within the originating institution throughout the study.

This provisioning step established the technical capability for swarm participation but did not grant access to sensitive data. Model training was initiated exclusively by an authorised researcher with approved access to the local data. All data preprocessing and model execution occurred locally, and patient-level data always remained confined to the institutional processing environment.

During training, each site trained the model on local data and participated in iterative parameter-synchronisation rounds with the swarm. Only model parameters (network weights) and scalar training metrics (loss values) were exchanged between institutions. Raw data and intermediate gradients were not shared. Parameter synchronisation frequency was configurable. In the experiments reported here model parameters were merged across the swarm every 10 local training epochs. Training proceeded either for a predefined number of epochs or until early-stopping criteria specified in the training configuration were met; the choice of stopping strategy did not affect the underlying execution model and is reported in the *Results* section.

## Results

### Deployment of a distributed learning infrastructure across university hospitals

The swarm learning architecture was successfully deployed across the three largest Finnish university hospitals: HUS, TYKS, and TAYS. Each institution operated as an independent federated node using the same containerised swarm learning architecture. The regulator-assessed deployment was implemented at HUS within the Findata-certified Acamedic secure processing environment, whereas partner sites deployed equivalent components within their local institutional processing environments under local governance procedures.

Deployment established a reusable federated-learning capability that combined local data control with decentralised model coordination. Operational responsibilities remained separated between infrastructure administration and authorised researchers, and trained model artefacts remained under institutional control throughout the workflow. Upon completion of training, all participating sites obtained an identical trained model while retaining established governance and output-release procedures. Deployment, including the distribution of approved model code, was achieved without modification of existing procedures for code import, data access, or result release, enabling federated learning to be introduced as a governed extension to an already operational secure processing environment.

Training operations were fully traceable through local training logs, container logs, and coordination-layer records. The distributed ledger maintained a shared record of participant registration, parameter synchronisation, and merge operations across participating institutions, enabling independent reconstruction of model provenance and federated training history without reliance on a central coordinating authority.

### Security assessment findings and certification outcome

The security assessment focused on the swarm learning extension rather than the previously certified Acamedic platform itself. Assessment activities included documentation review, interviews, demonstrations, and technical security testing (Supplementary Table 1).

The assessment identified one high-severity, two medium-severity, and two low-severity findings (Table 1). The identified findings reflected practical operational risks introduced by adding decentralised swarm learning functionality to an existing secure processing environment. The high-severity finding demonstrated a potential pathway for data exfiltration through unrestricted DNS queries originating from the swarm learning virtual machine. Medium-severity findings concerned the possibility of privilege escalation through container configuration and incomplete central collection of swarm learning operational logs. Additional low-severity observations related to malware scanning of container images and isolation of an auxiliary licensing component. All high- and medium-severity findings were remediated and independently verified. Following successful remediation, KPMG issued a certification report confirming compliance of the assessed extension with applicable Findata requirements.

**Table 1.** Principal Findings of the differential security assessment.

| Finding | Potential impact | Remediation outcome |
| --- | --- | --- |
| DNS exfiltration pathway (High) | Potential bypass of the intended data isolation boundary | DNS forwarding restricted to approved endpoints; verified during reassessment |
| Container privilege escalation (Medium) | Researchers could obtain root privileges on swarm learning virtual machine | Container permissions hardened and remediation verified |
| Incomplete central log collection (Medium) | Reduced monitoring coverage and incident visibility | Centralised SPIRE server and agent logging implemented and verified |
| Container malware scanning gap (Low) | Malicious binaries may not be detected before deployment | Managed through compensating controls and mitigation plan |
| License server isolation (Low) | Potential attack path through an auxiliary component | Managed through risk acceptance and compensating controls |

The evaluated governance model explicitly separated infrastructure certification from study-specific risk assessment (Table 2). Infrastructure controls, including access management, logging, monitoring, network isolation, and data-egress restrictions, were assessed through the certification process, whereas risks arising from specific data-model-output combinations remained subject to separate study-level review and approval procedures.

**Table 2.**
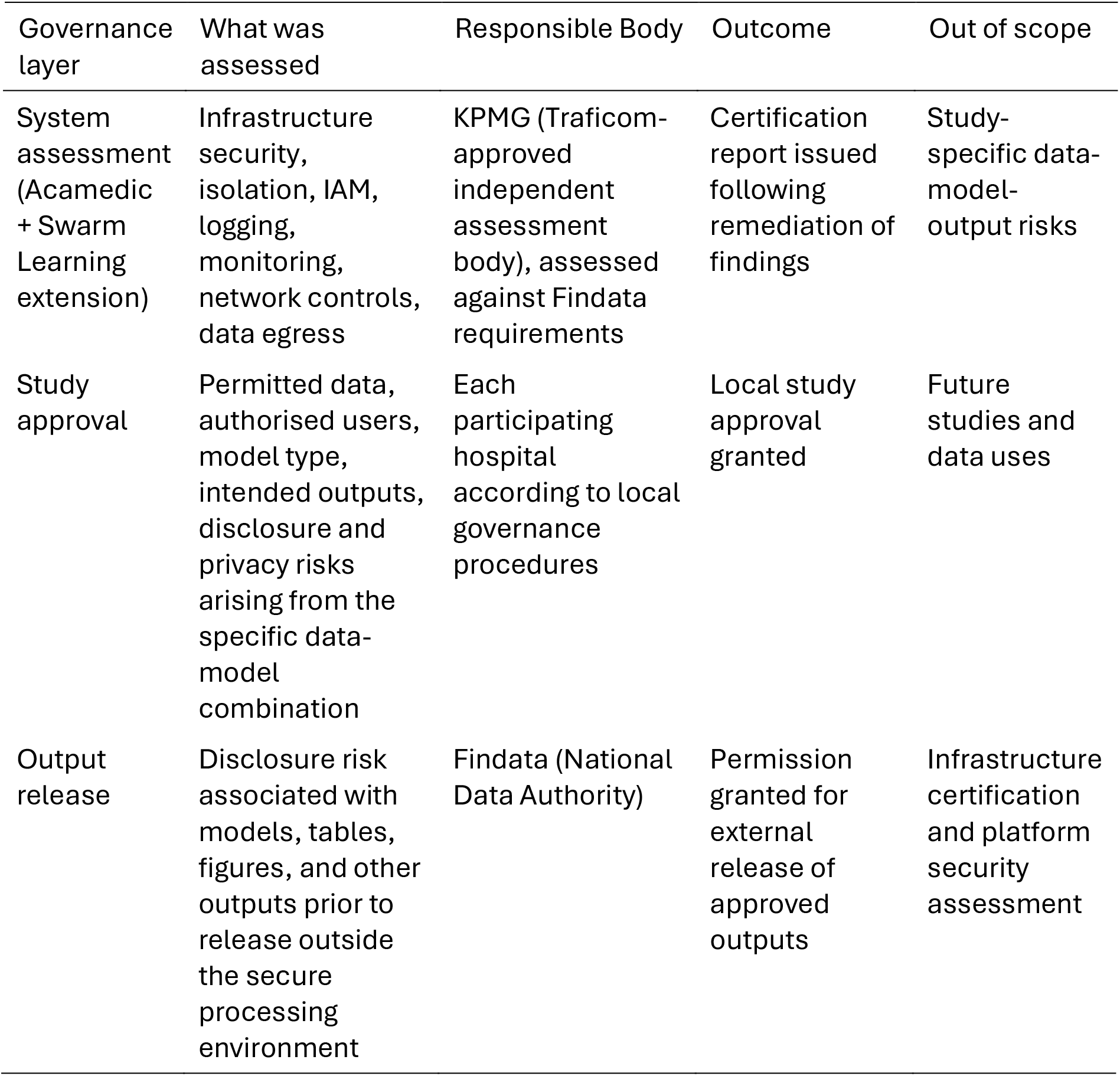
Governance model for infrastructure certification, study approval, and output release.

| Governance layer | What was assessed | Responsible Body | Outcome | Out of scope |
| --- | --- | --- | --- | --- |
| System assessment (Acamedic + Swarm Learning extension) | Infrastructure security, isolation, IAM, logging, monitoring, network controls, data egress | KPMG (Traficom-approved independent assessment body), assessed against Findata requirements | Certification report issued following remediation of findings | Study-specific data-model-output risks |
| Study approval | Permitted data, authorised users, model type, intended outputs, disclosure and privacy risks arising from the specific data-model combination | Each participating hospital according to local governance procedures | Local study approval granted | Future studies and data uses |
| Output release | Disclosure risk associated with models, tables, figures, and other outputs prior to release outside the secure processing environment | Findata (National Data Authority) | Permission granted for external release of approved outputs | Infrastructure certification and platform security assessment |

Following successful completion of the assessment, the swarm learning-enabled infrastructure was authorised for secondary use of health data under Finnish legislation and operationalised as a reusable extension to the HUS Acamedic secure processing environment for future permit-approved projects. The architecture was designed to align with key principles of emerging European secure processing environment frameworks, including data locality, controlled access, traceability, and auditable processing.^5,17,18^

### Operational exemplar: AML study population and data availability

The AML cohort included 2,240 adult patients treated at HUS (n=912), TYKS (n=689), and TAYS (n=639). Median age ranged from 60 to 66 years across participating hospitals, and median follow-up ranged from 359 to 529 days (Table 3).

**Table 3.**
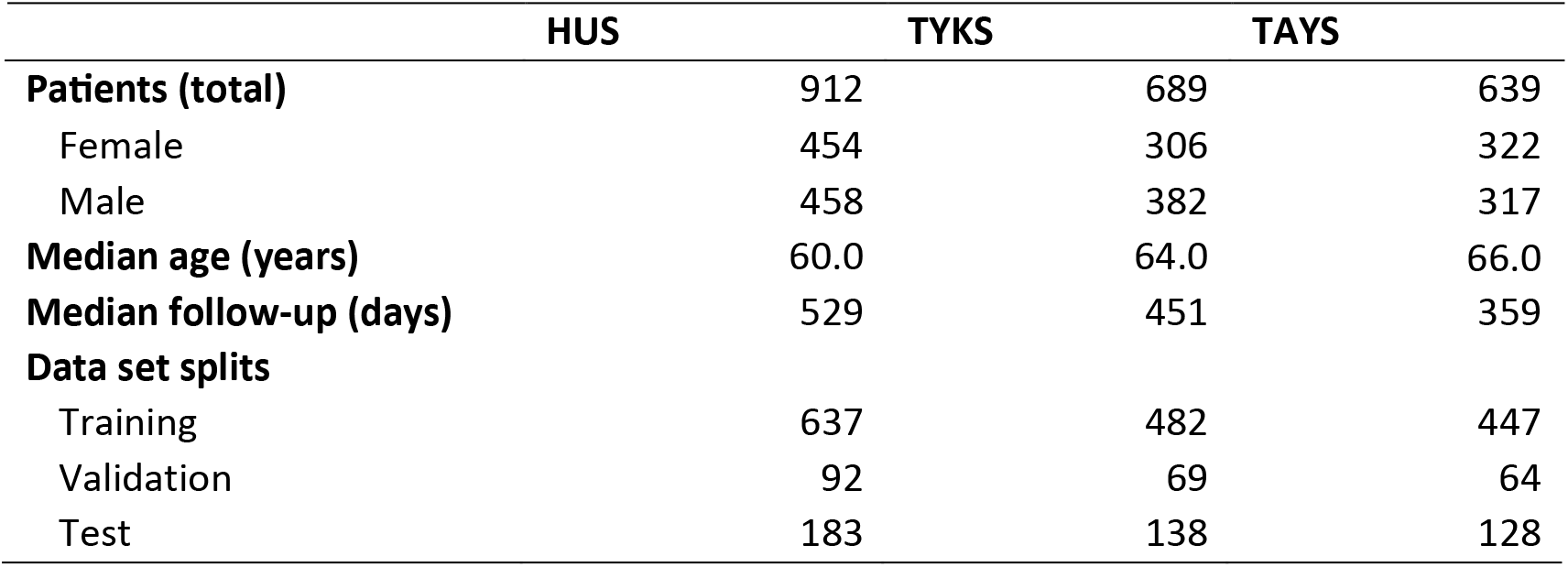
Cohort: Patient numbers of the AML cohort and basic demographic characteristics across the three sites.

Longitudinal laboratory measurements were available at all sites and included routinely collected blood counts, differential blood counts, lactate dehydrogenase, and peripheral blood blast measurements. Laboratory data were available at high temporal density within the predefined observation window surrounding AML diagnosis (Table 4).

**Table 4.** Model features and data availability across the three sites. Data is reported inside the study observation window: -5 to 14 days relative to AML diagnosis. *Average number of measurements per patient inside observation window. TYKS, Turku University Hospital; TAYS, Tampere University Hospital.

|  | **Patients with measurements** |  |  | **Measurements per patient\*** |  |  |
| --- | --- | --- | --- | --- | --- | --- |
|  | HUS | TYKS | TAYS | HUS | TYKS | TAYS |
| **Age at** |  |  |  |  |  |  |
| **Diagnosis** | 912 | 689 | 639 | - | - | - |
| **Hemoglobin** | 889 | 678 | 634 | 11.42 | 9.10 | 10.24 |
| **Leukocytes** | 889 | 678 | 634 | 11.42 | 9.10 | 10.22 |
| **Platelets** | 889 | 678 | 634 | 11.41 | 9.09 | 10.22 |
| **Lymphocytes** | 887 | 600 | 601 | 4.58 | 1.90 | 2.95 |
| **Monocytes** | 877 | 600 | 598 | 4.27 | 1.89 | 2.92 |
| **Neutrophils** | 882 | 228 | 197 | 4.48 | 0.44 | 1.01 |
| **Eosinophils** | 797 | 599 | 577 | 3.33 | 1.88 | 2.75 |
| **Basophils** | 744 | 598 | 559 | 2.92 | 1.88 | 2.68 |
| **LDH** | 844 | 545 | 502 | 4.85 | 3.34 | 1.96 |
| **Blasts (PB)** | 680 | 457 | 491 | 2.93 | 1.46 | 2.47 |

### Distributed training execution and operational performance

Swarm learning was successfully executed across all three university hospitals, with each institution operating as an independent federated node. No patient-level data or intermediate gradients were exchanged during training. Only encrypted model parameters and scalar training metrics were shared during synchronisation rounds, whereas all coordination events were recorded through the blockchain-based coordination layer.

Training and validation loss decreased throughout optimisation for both federated and local models. In the swarm learning setting, transient increases in loss were observed during scheduled parameter-synchronisation events; however, optimisation resumed immediately following each merge event (Figure 4A). Validation loss reached its minimum at epoch 40 during swarm training, compared with approximately epoch 22-25 for locally trained models (Suppl. Figure 1), indicating more sustained training without evidence of overfitting in the swarm setting. Parameter-merging operations completed successfully throughout execution, supporting the operational robustness of decentralised training across institutions.

**Figure 4.**
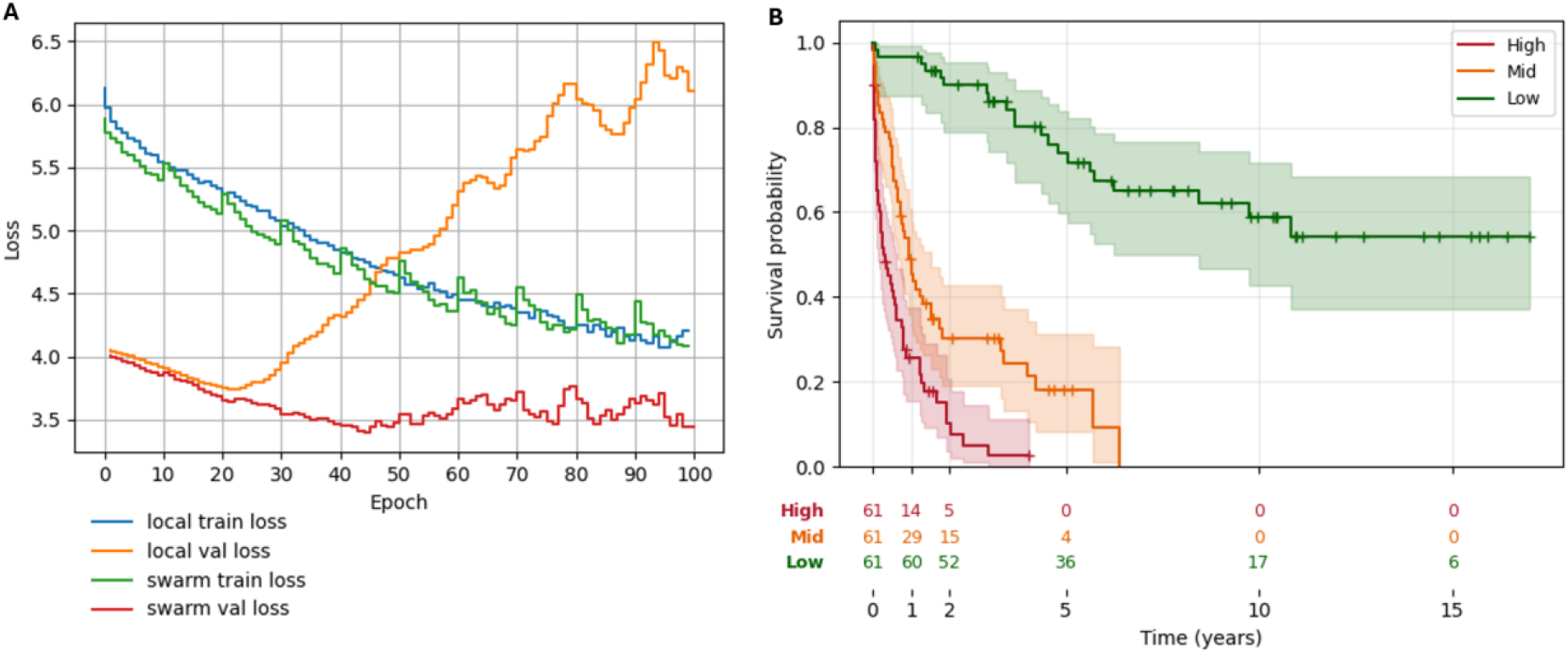
Swarm learning of an AML survival-prediction model as operational use-case. **(A)** Evolution of training and validation loss over training epochs for a federated acute myeloid leukaemia (AML) survival model. Swarm learning using all three participating hospital nodes (HUS, TYKS, TAYS). Curves show training loss and validation loss from HUS as a function of epoch. Vertical grey lines indicate swarm synchronization points at which model parameters were exchanged and merged across nodes (every 10 local training epochs in this experiment). The optimal validation loss from the swarm training run: epoch 40. **(B)** Model-based risk stratification on the independent test dataset from HUS for the swarm-trained model. Kaplan–Meier overall survival curves for patients stratified into high, medium, and low risk groups based on quantiles of the model predicted risk scores. Evaluation of the swarm-trained model used the model parameter from epoch 40, corresponding to the optimum identified during swarm training across all three hospitals.

### AML operational exemplar: model performance

To evaluate the clinical utility of the deployed infrastructure, we assessed the performance of federated AML survival models on independent hold-out test sets from each participating institution. Patients were stratified into prognostic risk groups based on model predicted risk scores and evaluated using Kaplan–Meier analysis.

Both federated and local models produced clinically meaningful separation between low-risk and high-risk patient groups. However, the swarm-trained model consistently achieved stronger risk separation across all three independent test cohorts (Suppl. Figure 2). Patients classified as high risk by the swarm-trained model exhibited substantially poorer survival outcomes than those classified as low risk (Figure 4B). Relative to locally trained models, the swarm-trained model exhibited larger hazard ratios, greater Kaplan– Meier curve separation, improved two-year survival discrimination, improved discrimination (C-index), and lower prediction error (integrated Brier score) (Supplementary Tables 3-5).

Together, these findings demonstrate that regulator-assessed federated learning can be operationalised across independent healthcare institutions while preserving data locality, maintaining auditable and traceable training operations, and producing clinically interpretable model outputs.

## Discussion

This study shows that federated learning can be integrated into a certified secure processing environment, subjected to differential regulatory assessment, and operationalised across independent university hospitals without centralising patient-level data. The principal contribution is a reusable infrastructure and governance model that separates system-level assessment of the federated extension from study-specific assessment of data, models, and outputs. Using AML survival modelling as an operational exemplar, the system supported stable decentralised training and prognostic risk stratification on independent test sets from all three participating hospitals. At HUS, swarm learning was integrated into the Findata-certified Acamedic SPE and underwent differential regulatory assessment. TYKS and TAYS implemented the same technical architecture within locally governed institutional environments.

Previous studies have demonstrated federated learning using real hospital data, but published evidence describing its assessment and operation within a formally regulated SPE remains limited.^15,1^ This translational gap reflects a broader challenge identified in a recent Nordic-Baltic federated health-data initiative, in which organisational coordination, divergent interpretations of privacy and risk, governance complexity, and the absence of harmonised legal frameworks proved more limiting than the underlying technical solutions themselves.^12^ Federated learning algorithms and software frameworks are increasingly mature; however, mechanisms for governing, auditing, and operationalising these technologies within real-world health-data environments remain comparatively underdeveloped. This study addresses that gap by reporting the technical integration, differential security assessment, governance model, and operational use of a federated-learning extension within a certified secure processing environment.

A central contribution of this study is the explicit separation of infrastructure-level governance from study-level governance. The differential assessment at HUS evaluated controls that can be certified independently of a specific analysis, including identity and access management, network isolation, logging, monitoring, environment protection, and data-egress controls. In contrast, disclosure risks arising from a particular combination of data, model, parameter exchange, and analytical outputs were assessed through established study-approval and output-release procedures. This layered governance model enables federated learning to be embedded within existing regulatory workflows rather than treated as a project-specific exception. As a result, the federated capability becomes part of the assessed infrastructure, whereas the risks associated with individual analyses continue to be evaluated on a case-by-case basis.

The assessment also provides practical insight into the controls most likely to require attention when introducing federated-learning capabilities into regulated research environments. Notably, the principal findings did not concern federated optimisation algorithms themselves, but rather infrastructure controls surrounding their operation, including data-egress controls, privilege management, and audit logging. This observation supports the view that barriers to operational deployment are increasingly infrastructural and organisational rather than algorithmic.

Traceability was provided through complementary logging layers. The SPE retained its established system and user logging controls, local model and container logs recorded execution within each institution, and the swarm coordination layer recorded participant registration, parameter exchanges, and merge events in a ledger distributed across participating nodes. The distributed ledger did not replace statutory logging of the SPE system and users, security assessment, or institutional accountability. Rather, it provided an additional tamper-resistant shared record from which the sequence of federated coordination events and model lineage could be reconstructed.

The deployment also provides an example of how federated learning can be incorporated into the emerging European framework for secondary use of health data. Many of the controls implemented in this study correspond to requirements being operationalised through the European Health Data Space (EHDS) and the TEHDAS2 specifications for secure processing environments, including processing within controlled environments, permit-based access, data minimisation, restricted egress, independent assessment, traceability, and institutional control of data.^16–18^ Notably, the Finnish secure processing environments and their governance models were developed within the same regulatory context that informed European SPE development, making the deployment directly relevant to ongoing EHDS implementation efforts. However, the present study evaluated a national deployment and did not assess interoperability across HealthData@EU, cross-border authorisation procedures, or conformity assessment under a harmonised European framework. The findings should therefore be interpreted as an implementation example demonstrating how core EHDS and TEHDAS2 principles can be operationalised in practice, rather than as evidence of formal EHDS compliance.

Importantly, several capabilities expected of authorised secure processing environments under the EHDS were implemented as reusable infrastructure controls rather than as study-specific procedures. These included permit-governed access, controlled software deployment, restricted data egress, independent security assessment, remediation of identified findings, and comprehensive logging of processing activities. The distributed-ledger coordination layer further enabled reconstruction of participant registration, parameter exchanges, model updates, and training lineage across organisations, providing an additional mechanism for provenance tracking and traceability. Such capabilities are increasingly relevant beyond secondary use of health data. Under the EU AI Act, many health-related AI applications may be classified as high-risk systems and are therefore subject to requirements relating to record keeping, traceability, monitoring, and accountability.^4^ Similar expectations are reflected in Software as a Medical Device and In Vitro Diagnostics regulatory frameworks, where documentation of model provenance, system behaviour, and lifecycle management are central elements of governance.^20^ Although the present study did not evaluate conformity with these frameworks, it illustrates how federated-learning infrastructures can provide technical foundations that support future requirements for auditable and accountable AI in healthcare.

Operationally, the study showed that federated learning could be introduced through established procedures for software import, data access, execution, and result release rather than through a parallel governance pathway. This is important for reuse: the assessed capability can support subsequent permit-approved analyses without redesigning the underlying SPE, although each participating institution remains responsible for its local environment, approvals, and operational controls.

The AML analysis served as an operational test of whether the infrastructure could support an end-to-end modelling workflow using sensitive, longitudinal hospital data. The swarm-trained model completed training with 100% parameter-merge success and produced prognostic risk scores that separated survival outcomes on independent test sets from all three hospitals. In the reported run, separation between predicted risk groups and overall discrimination and prediction-error metrics were more favourable for the swarm model than for the corresponding local models. Together with the observation that swarm learning sustained training for a greater number of epochs without evidence of overfitting, these findings suggest that exposure to a larger and more diverse distributed dataset improved model learning, consistent with a recent systematic review reporting superior performance of decentralised compared with local learning.^21^ Because the analysis used a single modelling configuration, lacked a pooled-data benchmark, and was not compared with established clinical prognostic systems, it should be interpreted as evidence of analytical utility of the infrastructure rather than clinical utility of the AML model.

Federated-learning frameworks differ in orchestration, trust assumptions, governance support, and privacy mechanisms. Frameworks such as Flower and NVIDIA FLARE commonly support server-coordinated training, whereas Vantage6 and the Personal Health Train emphasize governed distributed analytics.^9,22–24^ Swarm learning was selected here because its peer-to-peer coordination and distributed ledger were compatible with the intended multi-institutional trust model.^10^ The study does not establish that swarm learning is generally superior to alternative federated-learning architectures.^21^ Rather, it demonstrates that a decentralised federated-learning design, operating without a central coordinating server and using distributed coordination between participating institutions, can be integrated into a certified secure processing environment and governed with existing regulatory frameworks.

The study has three principal strengths. First, it involved independent real-world university-hospital environments and routinely collected data rather than simulated federated partitions or curated research environments. Second, governance and deployability were evaluated as primary outcomes, including differential assessment and remediation of the HUS SPE extension. Third, the study distinguished infrastructure-level controls, study-specific data-model-output risks, and model-performance evaluation, providing a governance structure that can be reused beyond the AML exemplar.

Several limitations should be considered. First, regulatory assessment was performed only for the HUS deployment operating within the Findata-certified secure processing environment; although the same technical architecture was used at participating hospitals, governance and oversight at partner sites followed local institutional procedures. Consequently, the findings should be interpreted as demonstrating one operational model for integrating federated learning into secure processing environment governance. Second, the operational exemplar was limited to a single AML survival prediction use case across three university hospitals. While swarm learning outperformed local training in this setting, in line with evidence from a recent systematic review, the findings should not be interpreted as demonstrating superior performance of federated learning across all modelling tasks, data domains, or deployment contexts.^21^ Third, the study did not compare swarm learning with alternative federated optimisation strategies, privacy-enhancing technologies, or centralised pooled-data training under identical conditions. Finally, potential disclosure risks associated with trained model parameters were not evaluated experimentally in this study. Nevertheless, risks arising from the specific data-model-output combination were assessed through established study approval procedures. Consistent with Findata guidance and emerging EHDS and TEHDAS2 frameworks, such risks are context-dependent and should be evaluated in relation to the underlying data, model outputs, and intended use of the resulting models.^25^ Furthermore, although the distributed-ledger coordination layer supported traceability and reconstruction of training activities, its presence does not in itself demonstrate the absence of privacy leakage or establish compliance with regulatory frameworks governing high-risk AI systems or medical-device software.^4,20^

In conclusion, this study shows that federated learning can be integrated into an established secure processing environment, subjected to differential regulatory assessment, and operated across independent hospitals without centralising patient-level data. The principal contribution is a reusable governance and infrastructure model that separates assessment of system-level controls from study-specific evaluation of data, models, parameter exchanges, and outputs. The AML exemplar demonstrated that this infrastructure supports stable multi-site training and prognostic modelling on independent institutional datasets. More broadly, the findings illustrate a practical pathway for operationalising federated learning within regulated health-data environments and provide an implementation model relevant to emerging European frameworks for secondary use of health data.^5,17,18^

## Ethics approval

The study was conducted under the applicable data permits and institutional approvals at each participating hospital. The need for informed consent was assessed in accordance with Finnish legislation governing the secondary use of health and social data.

## Data availability

The patient-level data cannot be made publicly available because they contain sensitive health information and are subject to Finnish legislation and institutional data-permit conditions. Access may be requested from the relevant data controllers and authorities, subject to applicable approvals and secure processing requirements.

## Code availability

The data-preprocessing, model-training, and evaluation scripts will be made publicly available on GitHub upon final publication.

## Acknowledgements

We thank the technical, clinical, data-management, and information-security teams at the participating institutions for supporting the deployment and assessment of the federated-learning infrastructure.

## Supplementary material

### Data sources

Data were extracted locally from each site’s harmonized OMOP CDM instance. All source data originated from university hospital (tertiary care) electronic health records (EHRs). All sites used mappings shared through and developed collaboratively within the Finnish national OHDSI node, FinOMOP (www.ohdsi-europe.org/index.php/national-nodes/finland, github.com/FinOMOP/FinOMOP_mappings). To reduce re-identification risk, both extraction dates and database coverage are reported at calendar-year granularity.

#### 1. Data extraction

Data were extracted at all sites in 2025 using a Jupyter notebook developed and distributed by the coordinating centre (HUS). Eligible patients were identified based on a condition occurrence of acute myeloid leukaemia (AML), using OHDSI Standardized Vocabulary hierarchies^8^, OMOP standard concept 140352 “Acute myeloid leukaemia, disease” and all descendant concepts (https://athena.ohdsi.org/search-terms/terms/140352). The earliest AML condition occurrence available in the OMOP CDM was 2004 at HUS, 1986 at TAYS, and 2004 at TYKS. All sites contained AML condition records through the extraction date in 2025.

For all eligible patients, the following data were extracted: longitudinal laboratory measurements detailed in Table 2 within a window of −5 to +14 days relative to the initial diagnosis date; sex; year of birth; first and last diagnosis date; death date from the OMOP CDM death table; and date of the last recorded hospital visit from the OMOP CDM visit_occurrence table.

Age at diagnosis was derived from year of birth and year of first diagnosis, and only age at diagnosis was retained in the final extract. Overall survival (OS) was defined as the number of days from first diagnosis to death. For patients without a recorded death event, censoring time was defined as the number of days from first diagnosis to the end date of the last recorded hospital visit.

The concept set used for extraction was exported from OHDSI Atlas (github.com/ohdsi/atlas) and is available in the study GitHub repository [LINK].

#### 2. Cross-site data assessment

To assess data completeness and heterogeneity across participating sites, the extraction notebook generated aggregate summaries of patient counts, demographic characteristics, and feature availability, defined as non-missing measurement counts per patient within the prespecified extraction window.

The summaries were reviewed locally for re-identification risk before being shared with the coordinating site (HUS) for cross-site comparison. Following review, the extracts were transferred to each site’s swarm learning environment, where preprocessing and harmonization of variable names were performed as described in Section 4. At this stage, all dates had already been converted to values relative to diagnosis to minimize privacy risks.

The extraction notebook is available in the study GitHub repository [LINK].

#### 3. Cohort construction

The final analytical dataset was derived independently at each site from the extraction outputs. Patients aged <18 years at diagnosis were excluded.

A 14-day landmark approach was applied, retaining only patients who were alive and uncensored at day 14 following diagnosis. Individuals with an overall survival duration of ≤14 days were excluded irrespective of event status. All covariates were restricted to data available within the first 14 days following diagnosis, defining a post-landmark survival prediction task. The 14-day landmark was selected to capture the period of highest laboratory measurement density following AML diagnosis while keeping the observation window sufficiently short to support early risk stratification.

The cohort was subsequently partitioned into training (70%), validation (10%), and test (20%) sets using patient-level stratified sampling by survival status to maintain comparable event rates across partitions. Stratified sampling was performed independently within each site after cohort construction and landmark exclusions.

#### 4. Preprocessing of longitudinal laboratory data

First, all continuous longitudinal covariates underwent z-score standardization, with means and standard deviations estimated from the training partition. These parameters were subsequently applied to the validation and held-out test partitions.

Second, missing values were imputed using mean imputation on the standardized scale (constant value 0 after z-score standardization) using statistics derived from the training partition. Finally, binary missingness indicators were appended for all variables.

Preprocessing choices were informed by exploratory analyses performed on data from one participating site (TYKS) excluding the independent test set. These analyses compared alternative preprocessing strategies, including inclusion of missingness indicators and temporal downsampling, and informed selection of the preprocessing pipeline used in the reported experiments.

The study GitHub repository includes a notebook demonstrating the preprocessing pipeline using mock data.

### Model development

#### 5. Model architecture

The survival models were based on the DeepSurv framework, a deep neural network extension of the Cox proportional hazards model that optimizes the negative partial log-likelihood function.^26^ We used the implementation provided by the pycox package (github.com/havakv/pycox).^27^

The model architecture consisted of a two-layer fully connected neural network mapping 402 input features (demographics, laboratory measurement values, and binary missingness indicators) to a single scalar output representing the log-risk function. Each hidden layer contained 16 nodes with rectified linear unit (ReLU) activation, followed by batch normalization and dropout regularization (dropout rate 0.1).

#### 6. Model parameters

Training was performed for 100 epochs. Given the relatively modest cohort sizes, training was performed using a batch size equal to the full training partition. No early stopping was applied to observe complete training dynamics. Models were optimized using the Adam optimizer with a constant learning rate of 0.003. No learning-rate scheduling was applied. The reported architecture and hyperparameters (two hidden layers with 16 nodes per layer, batch normalization, and dropout rate 0.1) were selected based on exploratory analyses performed on training and validation partitions from one participating site (TYKS). No formal hyperparameter optimisation was performed. All other training parameters were kept at their default values as specified by torchtuples version 0.2.2 (github.com/havakv/torchtuples).

#### 7. Model selection

For both swarm-trained and locally trained models, the final model was selected as the checkpoint corresponding to the minimum validation loss on the respective site’s validation set.

Unless otherwise stated, all reported analyses correspond to the experiment using a data-split seed of 42.

#### 8. Local model training

For each swarm-trained model, a local comparator model was trained independently at each participating site using the same architecture, preprocessing pipeline, and hyperparameter settings as the corresponding swarm model.

The exact same training and validation partitions used for the corresponding swarm run were used for local training, but without parameter sharing or aggregation between institutions. Random seeds associated with model initialization and training were not fixed. Model selection was performed using minimum validation loss on the site’s validation partition, identical to the swarm training procedure.

#### 9. Model validation

The swarm model and its corresponding local comparator model selected through the validation procedure were evaluated on their respective held-out test partitions at each participating site.

Model discrimination was assessed using Harrell’s concordance index (C-index). Additional performance metrics are described in the corresponding Results section. Overall prediction error was assessed using the integrated Brier score (IBS).

For visualization of risk stratification, patients were grouped into tertiles according to model-predicted risk scores, with tertile thresholds determined independently within each test set. Kaplan-Meier survival curves were generated for each risk group using the *lifelines* package version 0.30.0 (github.com/camdavidsonpilon/lifelines) and estimated 2-year survival probabilities were reported.

Model calibration was assessed at 1, 2, and 5 years following diagnosis. Predicted survival probabilities at each time point were obtained from the model-predicted survival function. Patients were grouped into quintiles according to predicted survival probability at the corresponding evaluation time point. For each group, observed survival probabilities were estimated using the Kaplan-Meier method and compared with the corresponding mean predicted survival probabilities. Calibration plots report observed versus predicted survival probablility together with 95% confidence intervals for the observed Kaplan-Meier estimates.

#### 10. Code availability

The study GitHub repository at [LINK] contains the complete implementation code, Docker image specifications, package versions, and scripts required to reproduce the workflow. The repository additionally includes a minimal reproducible example based on mock data to facilitate validation of the preprocessing, training, and evaluation pipeline.

Reproducibility is limited by the fact that, although data-split seeds were retained, random seeds associated with model initialization and training were not retained. Consequently, exact numerical replication of individual model runs may not be possible, although the overall workflow remains reproducible.

**Supplementary Table 1.** Independent security assessment. Independent security assessment scope, methodology, and certification outcome

| Item | Description |
| --- | --- |
| Assessment trigger | Addition of swarm searning functionality to the previously certified Acamedic Secure Processing Environment |
| Regulatory framework | Findata Regulation 1/2022 and the Finnish Act on the Secondary Use of Health and Social Data |
| Assessment body | KPMG, a Traficom-approved independent information-security assessment body |
| Assessment target | Swarm learning extension; baseline SPE controls were not reassessed |
| Assessment methods | Documentation review, interviews, demonstrations, and technical security testing |
| Assessment period | August – December 2024 |
| Findings | 1 high, 2 medium, 2 low severity findings |
| Remediation | All high and medium findings remediated and verified |
| Outcome | Certification report issued for the assessed extension |

**Supplementary Table 2.** Assessment outcome details. Narrative summary of the independent delta security assessment of the HUS Acamedic swarm learning extension, including scope, evidence reviewed, principal findings, and remediation outcome

| **Audit domain** | **What was assessed in the delta security assessment** | **Evidence reviewed** | **Key findings** | **Outcome and remediation status** |
| --- | --- | --- | --- | --- |
| **Purpose and scope of the assessment** | Targeted reassessment of swarm learning functionality added to a previously certified secure processing environment (SPE) prior to production use | Documentation review, interviews, demonstrations, and technical testing | Focus restricted to new or modified controls introduced by the swarm learning extension. Not a full re-audit; controls previously assessed in the baseline SPE assessment were not reassessed | Assessment completed following verification of remediation of relevant high- and medium-severity findings |
| **1. Physical security** | No reassessment of physical security within the delta scope | Explicit exclusion in report | Not reassessed because covered in baseline SPE assessment and infrastructure hosted in a managed cloud environment | Not applicable within assessment scope |
| **3.1 Reliability of the operator** | Review of hosting arrangements and data residency (EU/EEA) relevant to operator reliability | Data protection impact assessment and environment security documentation | No new risks identified | Existing arrangements considered acceptable for the swarm learning extension |
| **3.2 Data protection** | Assessment of data protection design, DPIA coverage, and risk of data transfer outside the environment; confirmation that swarm learning does not share training data | DPIA documents, risk assessment materials, and contractual terms | No new risks identified; Architecture zero-trust by design and reflects data minimisation and non-sharing principles; | Controls considered appropriate for the assessed extension |
| **3.3 Premises** | No reassessment of physical premises within the delta scope | No new evidence generated | Not reassessed because covered in baseline SPE assessment | Not applicable within assessment scope |
| **3.4 Personnel** | Review of training, guidance, and user awareness for individuals accessing the environment | Training materials and onboarding documentation | No new risks identified; training and guidance processes reviewed | Personnel controls described as part of the existing control framework |
| **2.1 Identification (authentication and identity management)** | Assessment of user identification and authentication processes, including internal and external user flows and multi-factor authentication | Identity and access documentation and technical testing results | No new risks identified | Identification controls functioned as intended in tested scenarios |
| **2.2 User and access management** | Review of least-privilege access, workspace isolation, and lifecycle management of user permissions | Access governance and workspace management documentation | No risk identified; controls focus on role-based restriction and environment separation | Access controls considered adequate within the assessed extension |
| **2.3 Protecting the environment** | Assessment of technical protection measures, including network isolation, segmentation, CIS hardening, and defence-in-depth controls | Security configuration evidence, hardening benchmarks, and swarm learning technical documentation | Principal findings identified in this domain: ID01 (high): data exfiltration via DNS; ID02 (medium): privilege escalation via container configuration; ID04 (low): limited malware scanning in container workflow; ID05 (low): suboptimal component isolation for swarm learning license server | High- and medium-severity findings were remediated and verified; low-severity findings managed through compensating controls or accepted residual risk management |
| **2.4 Logs (logging and monitoring)** | Assessment of log collection, centralisation, retention, and monitoring | Logging plans and technical verification of log ingestion and monitoring | ID03 (medium): incomplete central log collection reduced monitoring coverage | Issue remediated, including centralisation and monitoring improvements; fix verified |
| **2.5 Management and monitoring** | Review of operational monitoring, alerting, and incident management processes | Incident handling procedures and monitoring documentation | No new risks identified; supports overall evaluation of operational security processes | Monitoring and response processes reviewed as part of the assessed extension |
| **Overall findings and certification implication** | Consolidated evaluation of technical findings identified during the delta security assessment | Findings summary (ID01–ID05) and remediation evidence | Five findings identified: one high, two medium, and two low severity, primarily related to environment isolation, privilege control, and logging | All high- and medium-severity findings were remediated and verified, and therefore did not prevent certification; low-severity findings managed as residual risk |

**Supplementary Table 3.**
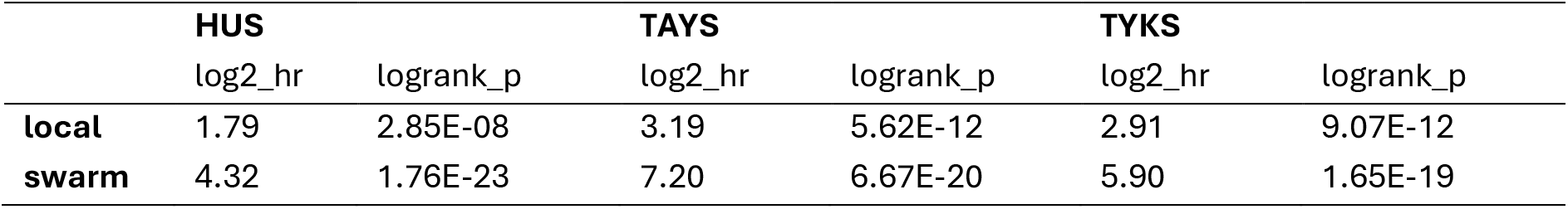
Model performance. Log hazard ratios and log-rank p values for locally trained and swarm-trained models evaluated on the independent test sets from each site. Shown are comparisons between the model-predicted high-risk and low-risk groups.

**Supplementary Table 4.**
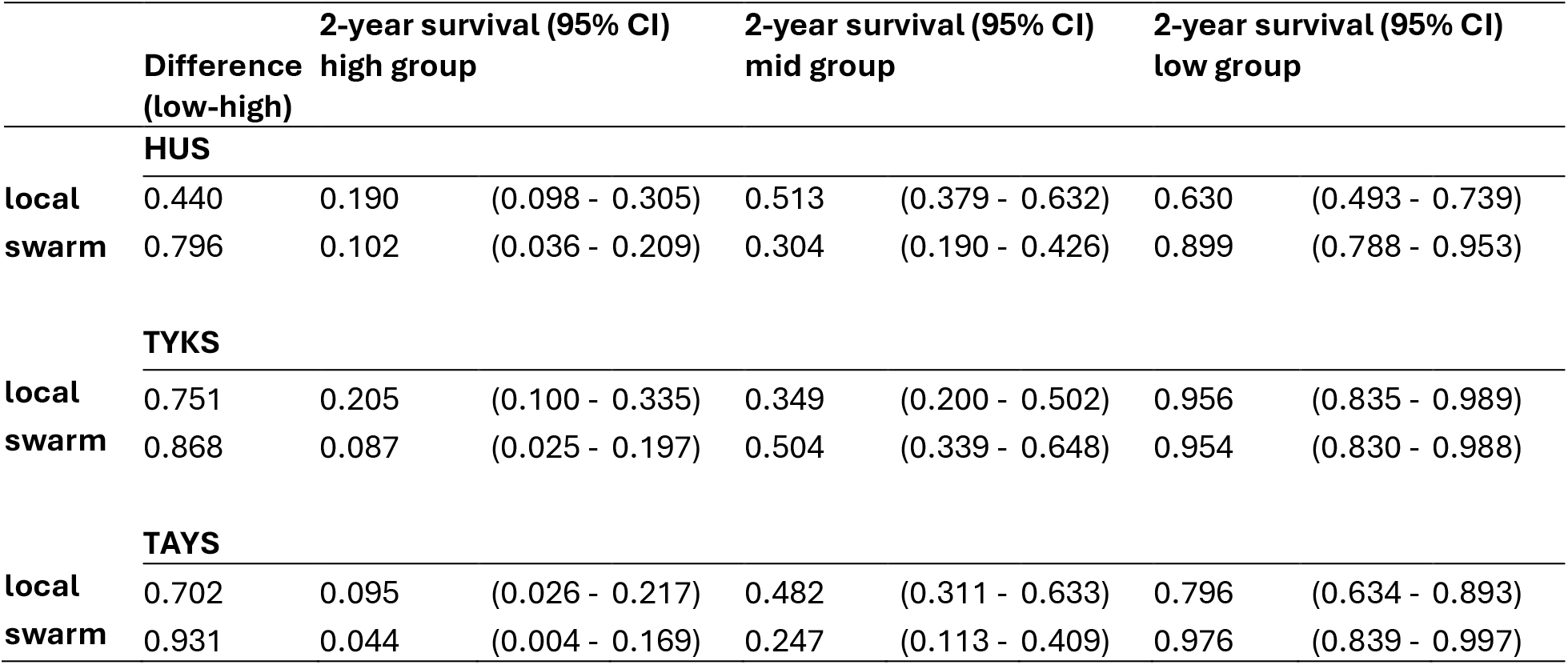
Model performance. Two-year survival estimates for model-predicted risk groups for locally trained and swarm-trained models evaluated on the independent test sets from the indicated sites. Difference denotes the absolute difference in two-year survival between the predicted high-risk and low-risk groups.

**Supplementary Table 5.** Model performance. Concordance index and integrated Brier score for locally trained and swarm-trained models evaluated on the independent test sets from each site. Higher concordance index indicates better discrimination, whereas lower integrated Brier score indicates better overall predictive accuracy.

|  | **C-index (higher better)** |  |  | **Brier (lower better)** |  |  |
| --- | --- | --- | --- | --- | --- | --- |
|  | HUS | TAYS | TYKS | HUS | TAYS | TYKS |
| **local** | 0.68 | 0.77 | 0.79 | 0.17 | 0.12 | 0.071 |
| **swarm** | 0.79 | 0.84 | 0.82 | 0.06 | 0.05 | 0.046 |

**Supplementary Figure 1.**
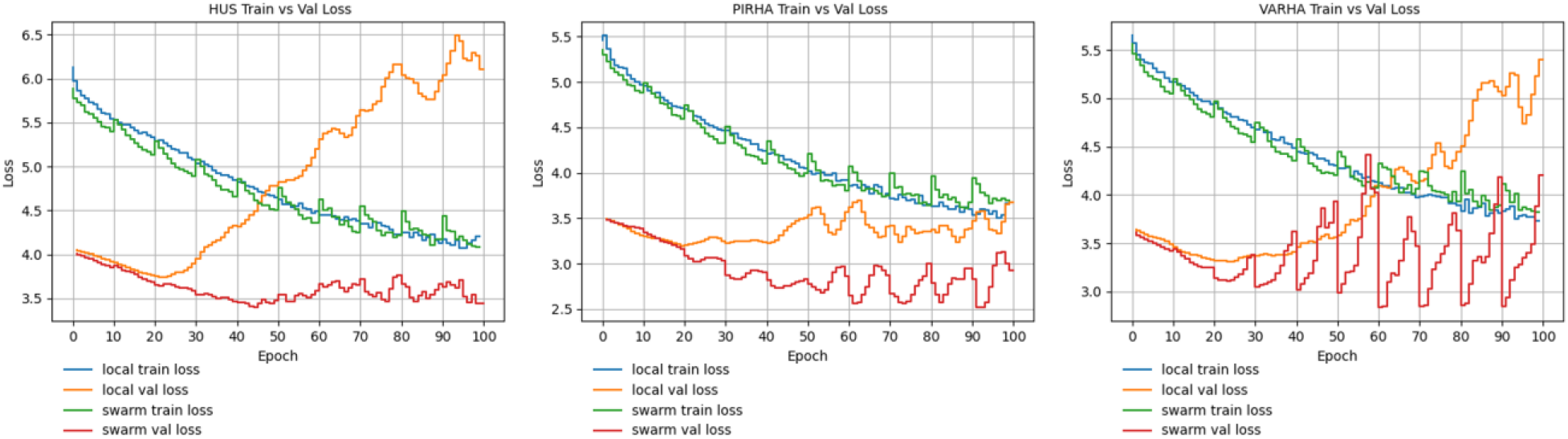
Evolution of training and validation loss over training epochs for all sites for the acute myeloid leukaemia (AML) survival model. Swarm learning using all three participating hospital nodes (HUS, TYKS, TAYS). Curves show training loss and validation loss from all three sites as indicated as a function of epoch. Vertical grey lines indicate swarm synchronization points at which model parameters were exchanged and merged across nodes (every 10 local training epochs in this experiment)

**Supplementary Figure 2.**
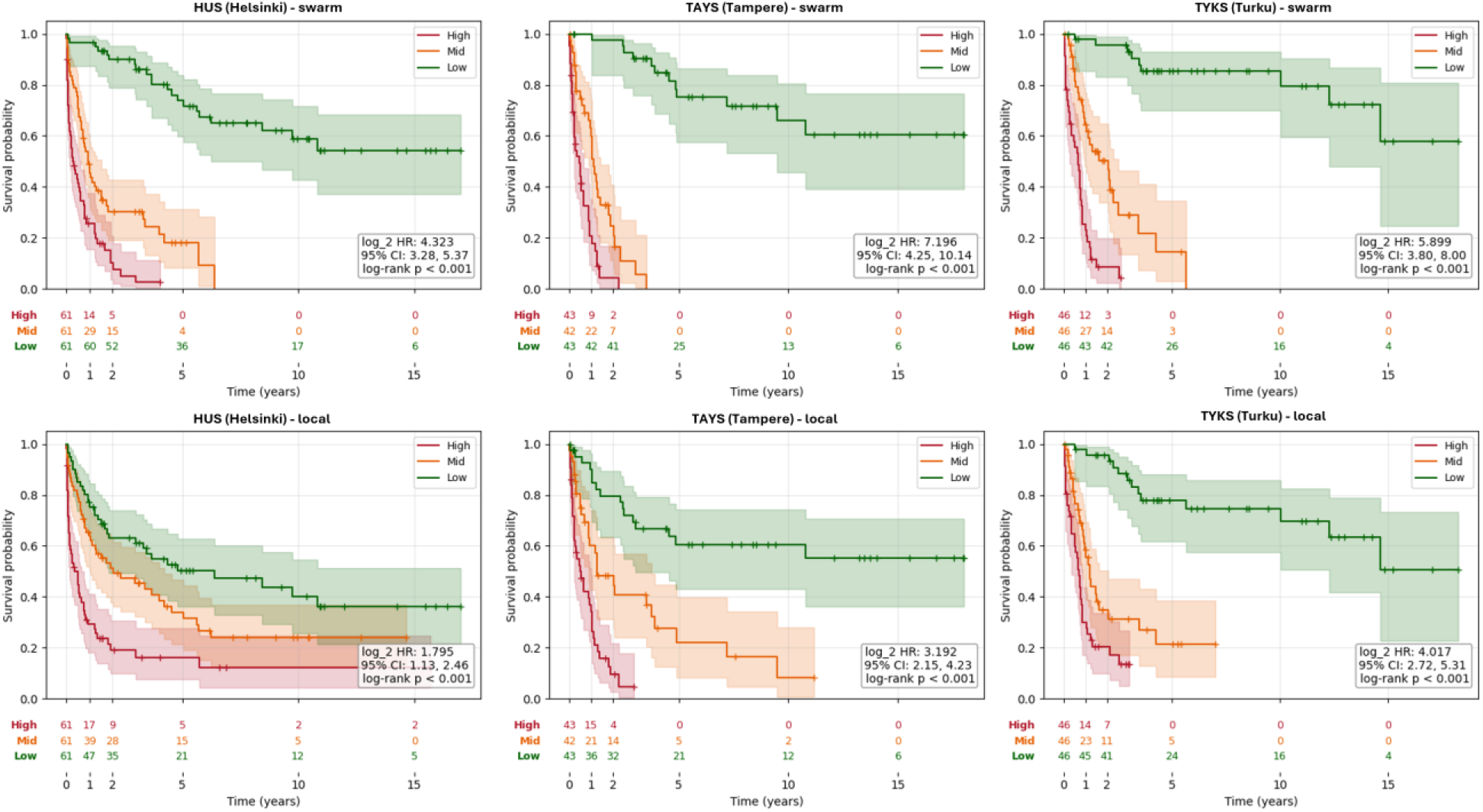
Model-based risk stratification on the independent AML test sets from each institution. Kaplan–Meier overall survival curves for patients stratified into high, medium, and low risk groups based on quantiles of the predicted risk scores from the trained model. Performances were evaluated on independent test sets from HUS, TAYS, and TYKS that were not used for model training, validation and model selection. Top row: Swarm-trained model evaluated at each site as indicated. Bottom row: Locally trained models. Evaluation of the swarm-trained model used the model parameter from epoch 40, corresponding to the global optimum identified during federated training across all three hospital nodes, see Figure 4. Patients at risk are shown below each plot.

## References

1. Azarfar, G. et al. Responsible adoption of multimodal artificial intelligence in health care: promises and challenges. *Lancet Digit*. Health 7, (2025).

2. Riley, R. D. et al. Importance of sample size on the quality and utility of AI-based prediction models for healthcare. *Lancet Digit*. Health 7, (2025).

3. Hernández-Sánchez, A. et al. Unravelling co-mutational patterns with prognostic implications in NPM1 mutated adult acute myeloid leukemia – a HARMONY study. Leukemia 40, 418–428 (2026).

4. EU AI Act. Regulation (EU) 2024/1c8S of the European Parliament and of the Council of 13 June 2024 Laying down Harmonised Rules on Artificial Intelligence and Amending Regulations (EC) No 300/2008, (EU) No 1c7/2013, (EU) No 1c8/2013, (EU) 2018/858, (EU) 2018/113S and (EU) 201S/2144 and Directives 2014/S0/EU, (EU) 201c/7S7 and (EU) 2020/1828 (Artificial Intelligence Act) (Text with EEA Relevance). (2024).

5. EHDS. Regulation (EU) 2025/327 on the European Health Data Space and Amending Directive 2011/24/EU and Regulation (EU) 2024/2847 - Public Health. https://eur-lex.europa.eu/eli/reg/2025/327 (2025).

6. HIPAA. Health Insurance Portability and Accountability Act of 1SSc. Pub. L. No. 104-1S1 1936 (1996).

7. Hripcsak, G. et al. Characterizing treatment pathways at scale using the OHDSI network. Proc. Natl. Acad. Sci. U. S. A. 113, 7329–7336 (2016).

8. Reich, C. et al. OHDSI Standardized Vocabularies—a large-scale centralized reference ontology for international data harmonization. J. Am. Med. Inform. Assoc. 31, 583–590 (2024).

9. Choudhury, A., et al. Advancing Privacy-Preserving Health Care Analytics and Implementation of the Personal Health Train: Federated Deep Learning Study. Jmir Ai 4, e60847 (2025).

10. Warnat-Herresthal, S. et al. Swarm Learning for decentralized and confidential clinical machine learning. Nature 594, 265–270 (2021).

11. Teo, Z. L. et al. Federated machine learning in healthcare: A systematic review on clinical applications and technical architecture. Cell Rep. Med. 5, 101419 (2024).

12. Peltonen, L.-M. C Chomutare, T. Federated learning’s uncomfortable truth: why human networks matter more than neural networks. J. Am. Med. Inform. Assoc. ocag047 (2026) doi:10.1093/jamia/ocag047.

13. Pati, S. et al. Federated learning enables big data for rare cancer boundary detection. Nat. Commun. 13, 7346 (2022).

14. Tang, R. et al. Pan-mediastinal neoplasm diagnosis via nationwide federated learning: a multicentre cohort study. *Lancet Digit*. Health 5, e560–e570 (2023).

15. Bakas, S., Li, X., Shah, P. C Roth, H. R. Federated Learning in Healthcare: From Research to Real-World Deployment. Annu. Rev. Biomed. Eng. 28, 163–186 (2026).

16. Raab, R. et al. Federated electronic health records for the European Health Data Space. *Lancet Digit*. Health 5, e840–e847 (2023).

17. Lodenius, H. et al. Guideline for data users on how to use data in a secure processing environment. https://doi.org/10.5281/zenodo.20157049 (2025) doi:10.5281/zenodo.20157049.

18. Lehväslaiho, H. et al. Technical specification for Health Data Access Bodies on the implementation of secure processing environments. https://doi.org/10.5281/zenodo.20266473 (2026) doi:10.5281/zenodo.20266473.

19. Mohammadi, M. et al. Differential privacy for medical deep learning: methods, tradeoffs, and deployment implications. Npj Digit. Med. 9, 93 (2026).

20. U.S. Food and Drug Administration. Software as a Medical Device (SaMD) - Good Machine Learning Practice for Medical Device Development: Guiding Principles. https://www.fda.gov/medical-devices/software-medical-device-samd/good-machine-learning-practice-medical-device-development-guiding-principles (2025).

21. Diniz, J. M. et al. Comparing decentralized machine learning and AI clinical models to local and centralized alternatives: a systematic review. Npj Digit. Med. 9, 174 (2026).

22. Moncada-Torres, A., Martin, F., Sieswerda, M., Van Soest, J. C Geleijnse, G. VANTAGE6: an open source priVAcy preserviNg federaTed leArninG infrastructurE for Secure Insight eXchange. AMIA. Annu. Symp. Proc. 2020, 870–877 (2021).

23. Beutel, D. J., et al. Flower: A Friendly Federated Learning Research Framework. Preprint at 10.48550/arXiv.2007.14390 (2022).

24. Roth, H. R., et al. NVIDIA FLARE: Federated Learning from Simulation to Real-World. https://doi.org/10.48550/arXiv.2210.13291 (2022) doi:10.48550/arXiv.2210.13291.

25. Findata. Producing anonymous results. Findata https://findata.fi/en/services-and-instructions/producing-anonymous-results/ (2026).

26. Katzman, J. L. et al. DeepSurv: personalized treatment recommender system using a Cox proportional hazards deep neural network. BMC Med. Res. Methodol. 18, 24 (2018).

27. Kvamme, H., Borgan, Ø. C Scheel, I. Time-to-Event Prediction with Neural Networks and Cox Regression. Preprint at 10.48550/arXiv.1907.00825 (2019).

